# Association of Anxiety with Uncinate Fasciculus Lesion Burden in Multiple Sclerosis

**DOI:** 10.1101/2024.10.08.24315108

**Authors:** Erica B. Baller, Audrey C. Luo, Matthew K. Schindler, Elena C. Cooper, Margaret K. Pecsok, Matthew C. Cieslak, Melissa Lynne Martin, Amit Bar-Or, Ameena Elahi, Christopher M. Perrone, Donovan Reid, Bailey C. Spangler, Theodore D. Satterthwaite, Russell T. Shinohara

## Abstract

**Importance:** Multiple sclerosis (MS) is an immune-mediated neurological disorder that affects 2.4 million people world-wide, and up to 60% experience anxiety.

**Objective:** We investigated how anxiety in MS is associated with white matter lesion burden in the uncinate fasciculus (UF).

**Design:** Retrospective case-control study of participants who received research-quality 3-tesla (3T) neuroimaging as part of MS clinical care from 2010-2018. Analyses were performed from June 1^st^ to September 30^th^, 2024.

**Setting:** Single-center academic medical specialty MS clinic.

**Participants:** Participants were identified from the electronic medical record. All participants were diagnosed by an MS specialist and completed research-quality MRI at 3T. After excluding participants with poor image quality, 372 were stratified into three groups which were balanced for age and sex: 1) MS without anxiety (MS+noA, n=99); 2) MS with mild anxiety (MS+mildA, n=249); and 3) MS with severe anxiety (MS+severeA, n=24).

**Exposure:** Anxiety diagnosis and anxiolytic medication.

**Main Outcome and Measure:** We first evaluated whether MS+severeA patients had greater lesion burden in the UF than MS+noA. Next, we examined whether increasing anxiety severity was associated with greater UF lesion burden. Generalized additive models were employed, with the burden of lesions (e.g. proportion of fascicle impacted) within the UF as the outcome measure and sex and spline of age as covariates.

**Results:** UF burden was higher in MS+severeA as compared to MS+noA (T=2.02, P=0.045, Cohen’s *f*^2^=0.19). A dose-response effect was also found, where higher mean UF burden was associated with higher anxiety severity (T=2.08, P=0.038, Cohen’s *f*^2^=0.10).

**Conclusions and Relevance:** We demonstrate that overall lesion burden in UF was associated with the presence and severity of anxiety in patients with MS. Future studies linking white matter lesion burden in UF with treatment prognosis are warranted.

**KEY POINTS:** *Question:* Are white matter lesions that impact the uncinate fasciculus (UF) associated with anxiety in patients with multiple sclerosis (MS)?

*Findings:* This retrospective, case-control study of 372 patients with MS included 3 anxiety severity groups: 1) MS without anxiety (MS+noA, n=99); 2) MS with mild anxiety (MS+mildA, n=249); and 3) MS with severe anxiety (MS+severeA, n=24). We identified associations between anxiety and UF lesion burden. Specifically, we showed that MS+severeA had higher UF lesion burden than MS+noA, and worsening anxiety severity increased with greater UF burden.

*Meaning:* Lesion burden in the UF may contribute to anxiety comorbidity in MS.

## INTRODUCTION

Multiple sclerosis (MS) is an immune-mediated neurological disorder that affects 2.4 million people worldwide^1–3^. The rates of anxiety in MS are three times as high as the general population, with up to 60% of persons with MS experiencing anxiety symptoms over their lifetime^4^. Though anxiety in MS is associated with substantial disability, including worse physical functioning and quality of life^4–6^, the mechanisms of anxiety in MS are not well-understood^5^. The few studies that have attempted to relate neuroinflammation and white matter pathology to anxiety in MS have not identified a biomarker that directly links anxiety symptoms to brain pathology, leading some scientists to speculate that anxiety is a consequence of living with an unpredictable illness^7–10^. However, 40% of those with MS do not have anxiety, suggesting neural pathophysiology may contribute to anxiety risk. Here, we evaluated whether lesions affecting white matter tracts that connect brain regions known to be associated with anxiety circuitry contribute to anxiety in MS.

Preclinical studies in rodents and primates have consistently demonstrated the importance of the orbitofrontal cortex (OFC) and the amygdala in anxiety and stress circuitry^11–14^. Human neuroimaging studies have also shown convergent results^15,16^. Across anxiety disorders, patients with anxiety show low OFC activation and high amygdala activation in functional magnetic resonance imaging^17–19^. A recent study of anxiety in MS has also identified abnormalities in resting state activity in the prefrontal cortex and amygdala^20^. The prevailing hypothesis is that the OFC provides top-down control over the amygdala in healthy brain states, and disruption of that relationship leads to heightened amygdala activity, increased fear response, and subsequent activation of the sympathetic nervous system^18,21^. The OFC and amygdala are structurally connected via the uncinate fasciculus (UF), and this structural relationship is conserved across species^22,23^. Though lesions to the UF should theoretically lead to OFC/amygdala decoupling and cause anxiety, it is not possible to prospectively lesion the UF to test this hypothesis in humans.

Scientists have attempted to address this limitation by using diffusion imaging to study white matter variability between people with and without anxiety, but findings have been inconsistent. Though some studies have described associations between anxiety disorders and decreased fractional anisotropy in fascicles connecting the anterior and posterior cingulate, amygdala, and prefrontal cortex^17,24–26^, others have not identified these associations^27^. Additionally, nearly all studies exclude participants with brain diseases and therefore cannot be extrapolated to MS. To advance our understanding of the neuroanatomic vulnerability to anxiety in MS, as well as to better characterize the pathophysiology of anxiety, studies with large sample sizes that link lesion location and burden to anxiety are necessary.

Here, we assessed the relationship of anxiety to white matter lesion burden in the UF in a large sample of patients with MS who received research-quality structural imaging as part of routine care. We hypothesized that patients with a severe anxiety phenotype would have higher UF lesion burden than non-anxious MS comparators, and that anxiety severity would be associated with worse UF disease.

## METHODS

### Participants

Participants with MS were identified from the electronic medical record (EMR) via the University of Pennsylvania Data Analytic Center, including demographics, ICD-10 diagnoses^28^, medications, and depression screens including Patient Health Questionnaires (2– and 9-question forms [PHQ-2/9])^29^. The University of Pennsylvania Institutional Review Board approved this study.

### MS Diagnosis

Participants aged 18 and older were included if they received an ICD-10 MS diagnosis (G35) from a specialist in the Penn Comprehensive MS Center and received 3T MRI under the University of Pennsylvania MS protocol^30^. Because no current biomarkers reliably distinguish between MS subtypes and predict progression, all MS subtypes were considered together^31^.

### Anxiety Diagnosis and Severity Stratification

It is well-known that anxiety is underdiagnosed and undertreated in medical populations, which creates challenges for capturing anxiety phenotypes from the EMR^32–34^. In response, we developed an anxiety stratification method that integrated EMR data from three data fields: ICD-10 diagnoses, medication prescriptions, and mood symptom ratings (**Figure 1**).

**Figure 1.**
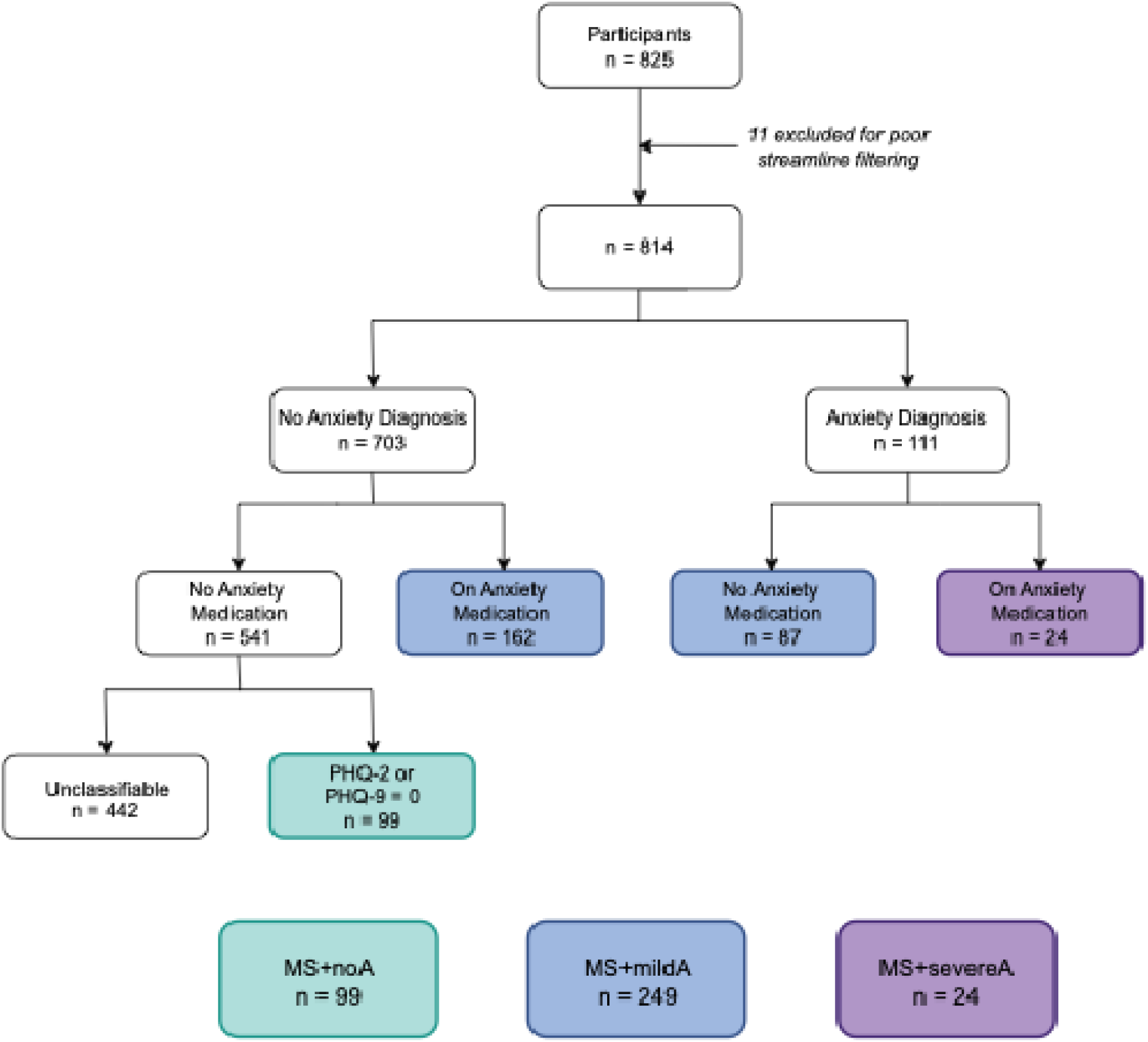
Flowchart of study population phenotyping. After excluding participants with poor-quality scans, patients with multiple sclerosis (MS) were stratified by anxiety diagnosis, prescription for anxiolytic medications, and psychiatric symptom screening. MS+noA, MS without anxiety; MS+mildA, MS with mild anxiety (anxiety diagnosis *or* on an anxiolytic medication); MS+severeA, MS with severe anxiety (anxiety diagnosis *and* anxiolytic medication); PHQ, Patient Health Questionnaire (2– or 9-question version).

Participants with MS were stratified into three groups, balanced for age and sex: 1) MS without anxiety (MS+noA); 2) MS with mild anxiety (MS+mildA); and 3) MS with severe anxiety (MS+severeA). MS+noA included persons who had no psychiatric diagnoses, took no psychiatric medications, and were asymptomatic on PHQ 2/9 (score=0). To classify MS+mildA and MS+severeA, we made two assumptions based on known provider undercoding behavior^35–37^. Specifically, we assumed that the presence of either an anxiety diagnosis (F40*) *or* a medication suggested some anxiety symptom burden (MS+mildA), whereas the presence of *both* an anxiety diagnosis and a medication prescription indicated that psychopathology was severe enough to be recognized and treated pharmacologically (MS+severeA; **Supplementary Table 1**)^38^. Participants without psychiatric diagnoses or medications were excluded from MS+noA if no PHQ-2/ 9 were available to confirm the absence of psychopathology. For all analyses, we defined “anxiety diagnosis” as a comparison between asymptomatic individuals and those with severe anxiety (MS+noA versus MS+severeA), and “anxiety severity” as a dose-response comparison across all groups (MS+noA, MS+mildA, and MS+severeA).

### PROMIS Validation

Given the assumptions required for group stratification, we next tested whether our anxiety diagnosis and severity stratification captured phenotypic variation in functioning. To do this, we evaluated group differences in Patient Reported Outcomes Measurement Information System (PROMIS) scores in a subset of patients, which has been validated in MS^39,40^. The PROMIS assesses current symptom burden in 10 domains, including mental health and mood, emotional problems, quality of life, physical health, social activities satisfaction, carrying out social activities, carrying out physical activities, fatigue, overall physical impairment, and overall mental impairment. Since the PROMIS was not used for anxiety stratification, it served as an independent measure of our phenotyping algorithm. For patients with multiple PROMIS scores, the score most proximal to their MRI was used.

We next calculated two summary measures, a Physical Functioning score (mean of PROMIS scores from physical health, carrying out physical activities, fatigue, and overall physical impairment) and an Emotional Functioning score (mean of PROMIS scores from mental health and mood, emotional problems, quality of life, social activities satisfaction, carrying out social activities, and overall mental impairment). Quality of life was included in Emotional Functioning given prior work suggesting that it better correlates with psychiatric versus physical impairment^41–44^. Finally, we tested whether our EMR-derived anxiety diagnosis and severity phenotyping were differentially related to Physical and Emotional Functioning. Multiple comparisons were accounted for by controlling the False Discovery Rate (Q<0.05).

### Image acquisition and processing

Structural 3T MRI was obtained as part of routine care using a research-quality protocol, including 3D T1w MPRAGE (TR=1.9s, TE=2.48ms, TI=900ms, FA=9°, acquisition time=4:18, 176 sagittal slices, resolution=1mm^3^) and 3D T2 FLAIR (TR=5s, TE=398ms, TI=1.8s, FA=120°, acquisition time=5:02, 160 sagittal slices, resolution=1mm^3^). Images were processed using a top-performing pipeline as previously described^30^. Briefly, T1w and FLAIR images were N4-bias corrected^45^, extracerebral voxels were removed from the T1w images using Multi Atlas Skull Stripping (MASS)^46^, T1w images and their corresponding brain masks were registered to the corresponding FLAIR, and skull-stripped FLAIR and aligned T1w images were intensity normalized using WhiteStripe^47^. MRI was usually acquired within 6 months of presentation to the MS clinic. For participants with multiple scans, their first clinical MRI was used.

### Automated Lesion Segmentation and Streamline Filtering

As previously described, we performed fully-automated lesion segmentation with the Method for Inter-Modal Segmentation Analysis (MIMoSA) to obtain binary maps of white matter lesions in a subject’s native space^48^. The quality of processed images and segmentations were assessed by a scientist with years of experience in MS imaging research.

To assess the association of white matter lesions to anxiety diagnosis and severity, we performed streamline filtering in DSI Studio^49–51^. We first identified the OFC and the amygdala as key brain regions relevant for anxiety and stress circuitry from preclinical and human neuroimaging research (**Figure 2A**)^16,18,21^. The UF, which structurally connects the OFC and amygdala, was then extracted from a standard atlas (**Figure 2B**)^50^. Next, individual lesion maps were normalized to the template space of the canonical UF (Montreal Neurological Institute 2009b Asymmetric template)^52^ using the T1-weighted–based transform calculated by antsRegistration (**Figure 2C**)^53,54^. Then, for both the right and left UF, streamlines intersecting lesions at any point in their trajectory were considered injured, isolated from the rest of the fascicle, and averaged (purple; **Figure 2D**). Finally, we created a summary measure of UF disease burden by dividing the mean volume of injured UF streamlines by the mean volume of streamlines in the canonical right and left UF (volume of injured streamlines/volume of healthy streamlines in canonical UF; **Figure 2E**).

**Figure 2.**
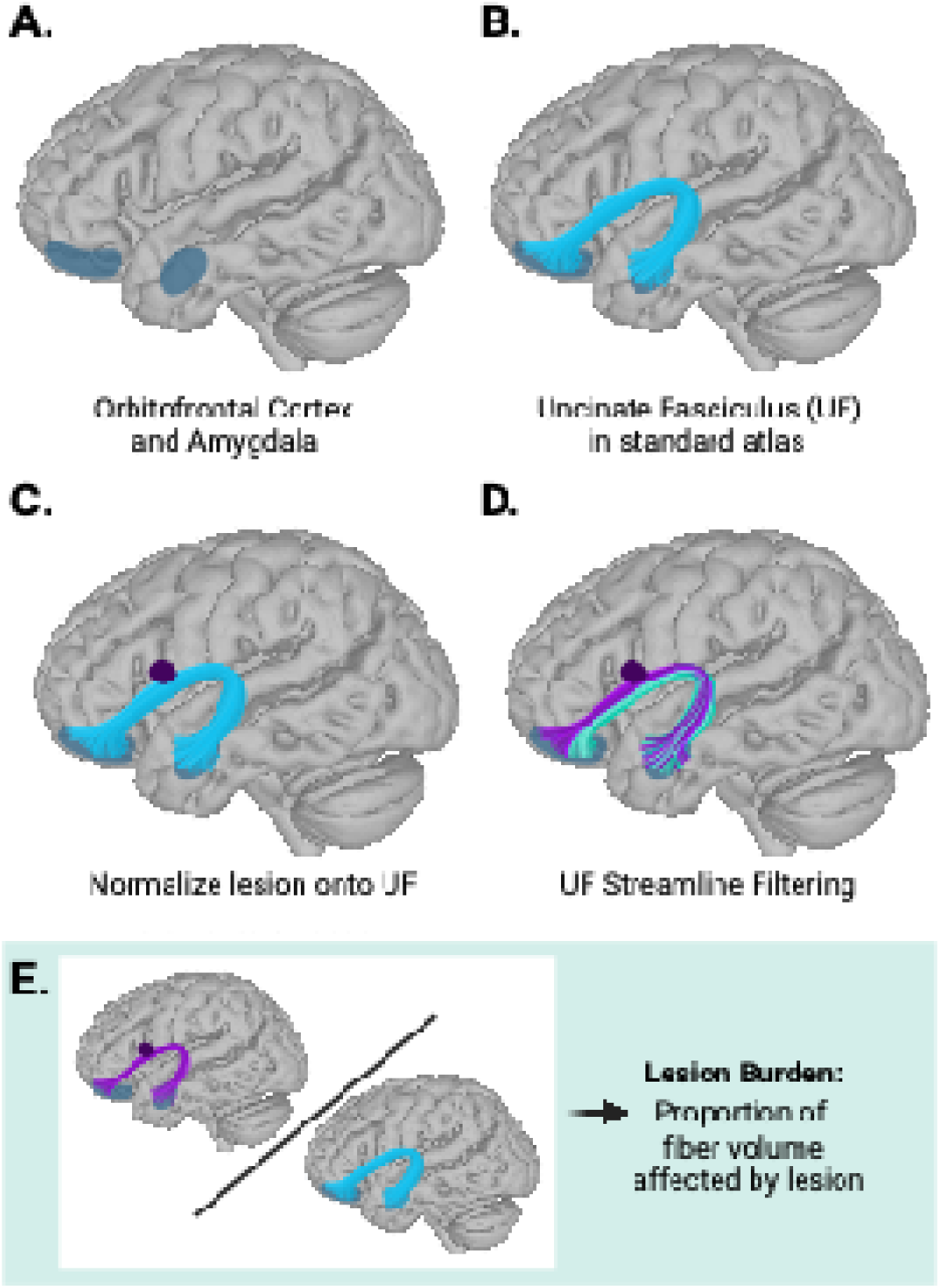
Uncinate fasciculus analysis pipeline. A) The orbitofrontal cortex (OFC) and the amygdala were identified as key brain regions relevant for anxiety and stress circuitry from preclinical and human neuroimaging research. B) The uncinate fasciculus (UF), which structurally connects the OFC and amygdala, was extracted from a standard atlas^50^. C) After automated white matter lesion segmentation with the Method for Inter-Modal Segmentation Analysis (MIMoSA), lesions were normalized to the template space of the canonical UF. D) For both right and left UF, streamlines intersecting lesions at any point in their trajectory were considered injured and isolated from the rest of the fascicle (purple). E) Lesion burden was defined as the ratio of the mean volume occupied by injured streamlines in UF divided by the total volume of streamlines in the canonical UF.

### Analysis of uncinate fasciculus

We first explored whether patients with an anxiety diagnosis had higher UF burden than those without psychiatric disease. To account for nonlinear changes in lesion burden with age, we used generalized additive models (GAMs; R-package mgcv^55^) to model mean UF burden between MS+severeA and MS+noA participants, covarying for sex and a spline of age. We then used GAMs to test for a dose-response relationship between greater UF burden and worse anxiety severity, modeling sex and a spline of age as covariates.

### Sensitivity Analyses

We selected the UF as a target fascicle of interest given its structural connections between the OFC and amygdala, which are part of a known anxiety circuit. However, there are other structural connections between the prefrontal cortex and the temporal lobe that are functionally connected but subserve different cognitive processes. For example, the fornix connects the prefrontal cortex and hippocampus and is involved in memory^56^. To test whether our results were specific to anxiety network white matter injury, we repeated our analyses but instead evaluated whether anxiety diagnosis and severity were associated with mean fornix burden.

Depression is frequently comorbid with anxiety in healthy populations and is present in up to 50% of patients with multiple sclerosis^30,57^. To test whether lesion burden in the UF was specific to anxiety severity or associated with general symptoms of both depression and anxiety, we stratified our group into samples with depression (MS+Depression) and psychiatrically asymptomatic (MS+noDepression; PHQ2/9=0, no psychiatric medications or diagnoses) using a previously described multi-step EMR phenotyping protocol^30^. We then tested whether UF burden was associated with a depression diagnosis.

### Code availability

Code and instructions for replicating all analyses can be found at: https://baller-lab.github.io/msanxiety/.

## RESULTS

After anxiety phenotype stratification, MS+noA included 99 persons (age[SD]=49.4[11.7], %female=75), MS+mildA included 249 persons (age[SD]=47.1[11.1], %female=82), and MS+severeA included 24 persons (age[SD]=47.0[12.2], %female=79; **Table 1**). Only seven people in the total sample of 372 participants (<2%) were prescribed steroids or interferon, which may cause psychiatric side effects.

**Table 1.** Participant demographics. All demographic variables were extracted from the electronic medical record. Race is patient-reported. P-values reflect ANOVA tests in continuous data (age, Patient Health Questionnaire-2) and chi-squared for categorical variables (sex, race, and MS medication). P-values for demographic differences between anxiety diagnosis (MS+noA vs MS+severeA) as well as P-values for anxiety severity (MS+noA, MS+mildA, MS+severeA) are displayed. MS, multiple sclerosis; SD, standard deviation; MS+noA, MS without anxiety; MS+mildA, MS with mild anxiety (anxiety diagnosis *or* anxiolytic medication); MS+severeA, MS with severe anxiety (anxiety diagnosis *and* anxiolytic medication); N.S., not significant; anti-CD20, medications that deplete CD20 B-cells (**Supplementary Table 2**).

|  | MS+noA | MS+mildA | MS+severeA | P<br>Anxiety<br>Diagnosis | P<br>Anxiety<br>Severity |
| --- | --- | --- | --- | --- | --- |
| n | 99 | 249 | 24 |  |  |
| Age (mean [SD]) | 49.44 [11.66] | 47.14 [11.14] | 46.96 [12.17] | N.S. | N.S. |
| Sex (% Male) | 25 (25.3%) | 46 (18.5%) | 5 (20.8%) | N.S. | N.S. |
| Race |  |  |  | N.S. | <0.001 |
| Asian | 2 (2.0%) | 0 (0%) | 0 (0%) |  |  |
| Black or African American | 19 (19.2%) | 54 (21.7%) | 3 (12.5%) |  |  |
| Patient Declined | 0 (0%) | 0 (0%) | 1 (4.2%) |  |  |
| Some Other Race | 10 (10.1%) | 7 (2.8%) | 1 (4.2%) |  |  |
| Unknown | 0 (0%) | 8 (3.2%) | 1 (4.2%) |  |  |
| White | 68 (68.7%) | 180 (72.3%) | 18 (75.0%) |  |  |
| MS Medication |  |  |  |  |  |
| On Anti-CD20 | 20 (20.2%) | 12 (4.8%) | 8 (33.3%) | N.S. | <0.001 |
| On Interferon | 0 (0%) | 0 (0%) | 1 (4.2%) | N.S. | 0.001 |
| On Steroids | 3 (3.0%) | 1 (0.4%) | 2 (8.3%) | N.S. | 0.006 |
| Patient Health Questionnaire-2 (mean [SD]) | 0.0 [0.0] | 0.57 [1.16] | 1.16 [1.54] | <0.001 | <0.001 |

### Anxiety is Differentially Associated with Physical and Emotional Functioning

A subset of participants also completed PROMIS scales (**Supplementary Table 3**). Across eight out of ten domains, MS+severeA had worse function than MS+noA, and worse anxiety severity was parametrically associated with worse functioning. We next compared the relationship of anxiety diagnosis and anxiety severity to our summary PROMIS scores (Physical Functioning and Emotional Functioning; **Figure 3)**. Higher anxiety severity was associated with worse functioning in both Physical and Emotional Functioning, with a larger effect size in the Emotional Functioning domain (Emotional Functioning: T=-4.2, P<0.001, Cohen’s *f*^2^*=*0.28; Physical Functioning: T=-2.5, P=0.015, Cohen’s *f*^2^*=*0.1).

**Figure 3.**
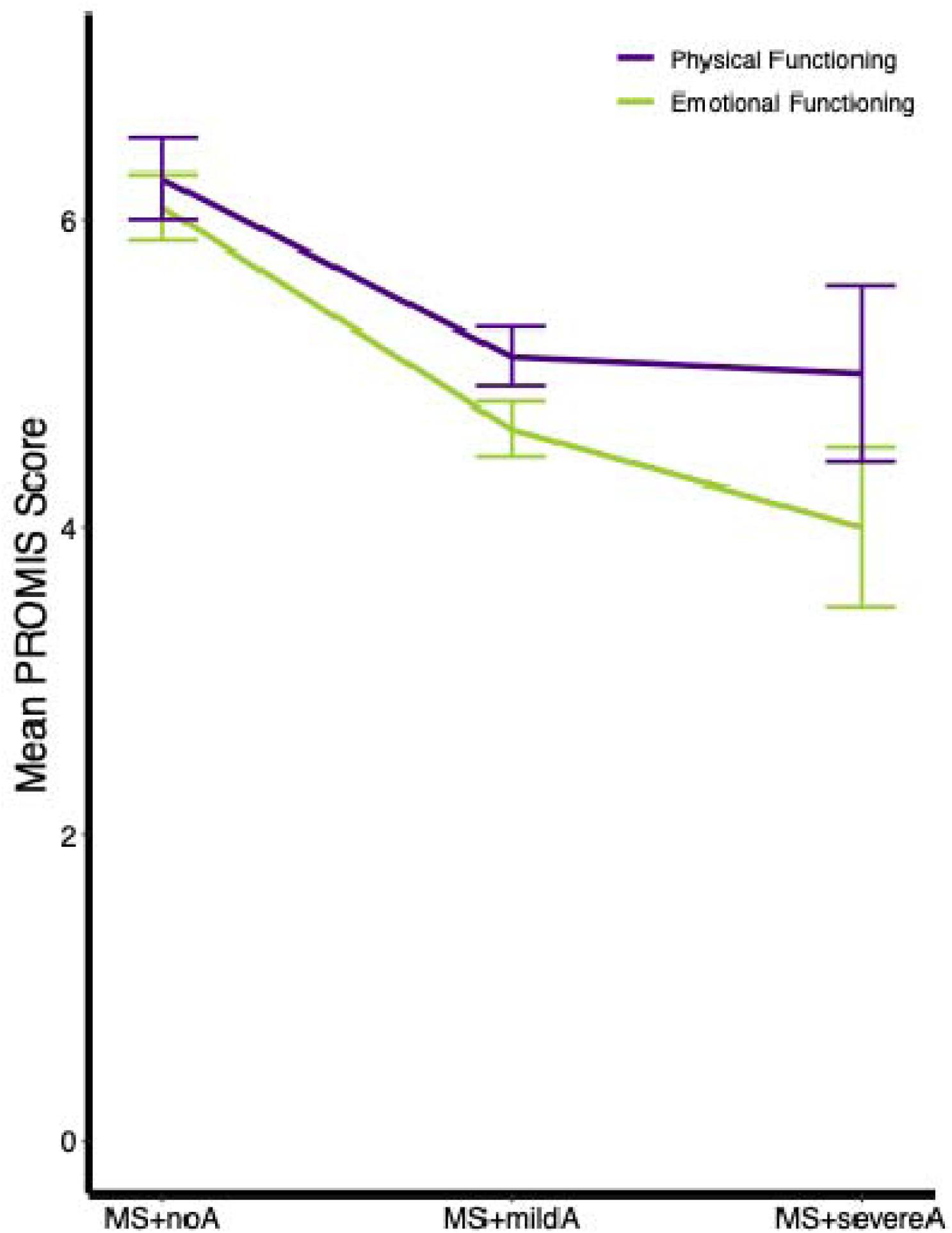
Patient-Reported Outcome Measures (PROMIS) decrease with higher anxiety severity. The PROMIS assesses current symptom burden in 10 domains, which were stratified into a Physical Functioning summary score and an Emotional Functioning summary score. Anxiety severity was associated with worse Physical (T=-2.5, P=0.015) and Emotional Functioning (T=-4.2, P<0.001; **Supplementary Table 4**). Post hoc pairwise t-tests also identified additional relationships: Emotional Functioning was worse in MS+severeA versus MS+noA (T=3.7, P*_fdr_*=0.015), and both MS+mildA and MS+severeA were associated with worse Emotional Functioning as compared with Physical Functioning in paired t-tests (MS+severeA: T=2.88, P*_fdr_*=0.047; MS+mildA: T=2.97, P*_fdr_*=0.015). Multiple comparisons were accounted for by controlling the False Discovery Rate (Q<0.05). Error bars reflect standard error of the mean.

Post hoc t-tests revealed that participants in the MS+severeA group had worse Emotional Functioning than MS+noA (T=3.7, P*_fdr_*=0.015, Cohen’s *d*=-1.87; **Supplementary Table 4**), but not the Physical Functioning domain (T=2.0, P*_fdr_*=N.S.). Additionally, participants in the MS+mildA and MS+severeA groups had worse Emotional versus Physical Functioning in paired t-tests (MS+severeA: T=2.88, P*_fdr_*=0.047, Cohen’s *d*=-0.69; MS+mildA: T=2.97, P*_fdr_*=0.015, Cohen’s *d*=-0.38), while there were no significant differences in MS+noA (T=0.68, P*_fdr_*=N.S.). These findings suggested that severe anxiety symptoms may be dissociable from the psychological impact of physical limitations.

### Anxiety Diagnosis and Severity are Associated with UF Lesion Burden

We next explored the relationship between anxiety and UF lesion burden. Anxiety diagnosis (MS+severeA vs MS+noA) was associated with UF lesion burden (T=2.02, P=0.045, Cohen’s *f*^2^=0.19; **Figure 4**). Additionally, we found a parametric relationship where greater UF lesion burden was associated with worse anxiety severity (T=2.08, P=0.038, Cohen’s *f*^2^=0.11).

**Figure 4.**
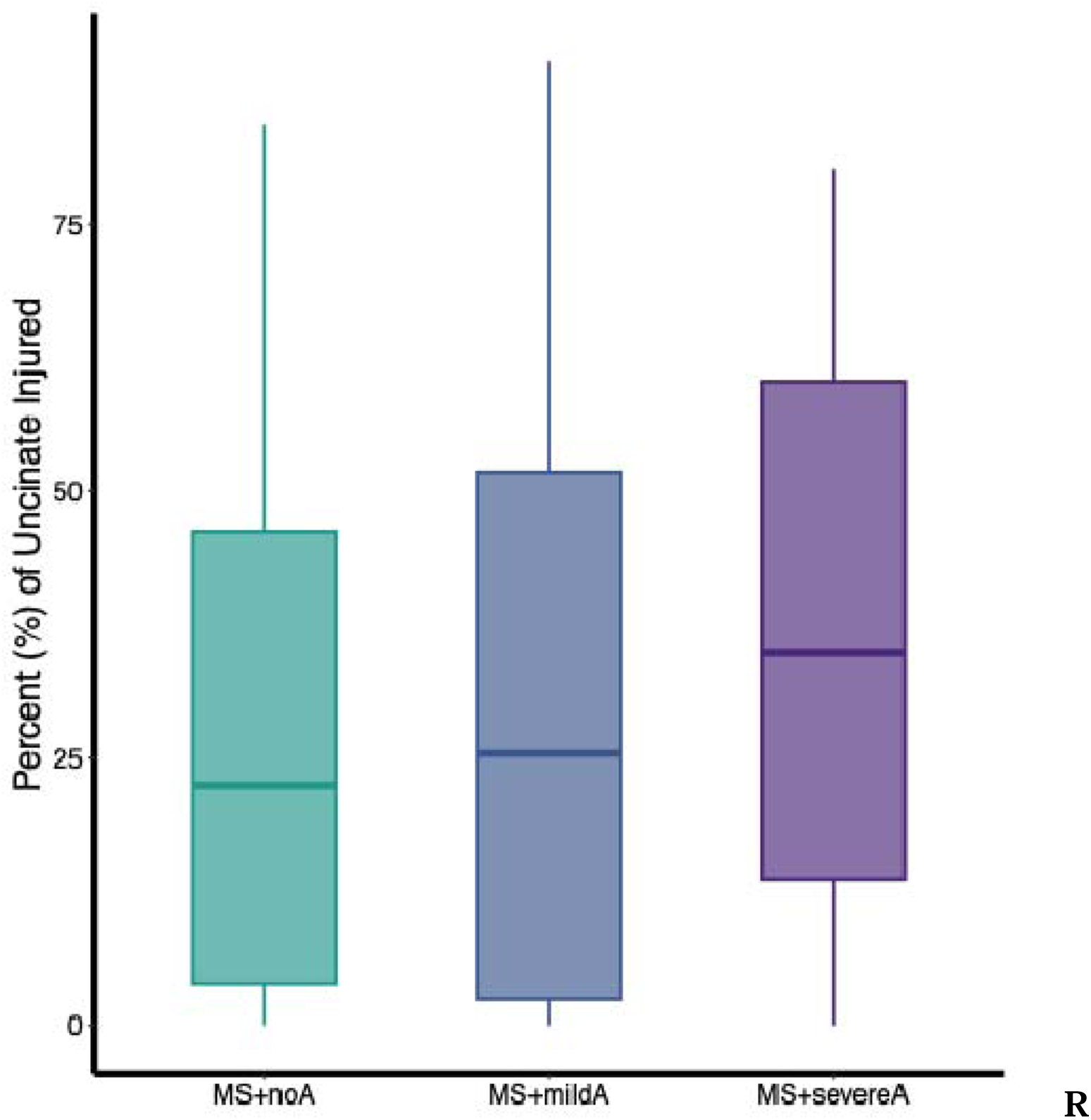
Higher uncinate fasciculus burden was associated with greater anxiety severity. Uncinate fasciculus (UF) lesion burden was higher in MS+severeA as compared to MS+noA (T=2.022, P=0.045). Across all three groups, greater UF lesion burden was also parametrically associated with worse anxiety severity (T=2.08, P=0.038).

### Anxiety-UF relationship is specific to both psychiatric phenotype and fascicle

To confirm that the relationship between anxiety and uncinate burden was specific, we first tested whether anxiety diagnosis or severity was associated with fornix injury, and neither analysis was significant (anxiety diagnosis: T=0.68, P=N.S.; anxiety severity: T=0.96, P=N.S.). We next compared UF lesion burden in EMR-derived depression phenotypes (MS+noDepression: n=99; MS+Depression: n=207). Again, no significant association was found (T=-1.7, P=N.S.).

## DISCUSSION

In this study, we developed an EMR phenotyping method to identify anxiety groups and derived a proxy measure of anxiety diagnosis and severity. We then demonstrated that anxiety in MS is associated with the burden of UF fasciculus lesions. Specifically, individuals with MS+severeA have a higher UF lesion burden than MS+noA. We also found a dose-response relationship, where greater anxiety severity was associated with increased UF burden. This relationship was specific to the UF rather than other prefrontal cortex-temporal white matter tracts and was specific to anxiety and not present in depression.

Previous studies have sought to disentangle the relationship between anxiety symptoms and MS^4^. One common hypothesis is that anxiety is a natural psychological consequence of living with a progressive neurological disorder and associated disability in physical and social functioning^9^. Others have hypothesized that MS neuropathology rather than physical disability alone may underlie anxiety severity. However, research parsing how lesion location or lesion volume is associated with anxiety have yielded inconsistent results^7^. In one prior study in 48 people with MS, anxiety was associated with white matter lesion load in the fornix^58^. Other studies have found no associations^9^. We attempt to address gaps in the literature by using a large sample of participants with research-quality imaging and coupling it with automated white matter lesion segmentation to decrease cost and time associated with image analysis. By evaluating lesion burden rather than lesion location or volume, we were able to identify a significant association between anxiety and disease in the UF. Our PROMIS results also suggested that physical functioning can be disentangled from emotional functioning in the MS+severeA group.

Our research adds to the growing body of literature showing that psychopathology in MS is related to injury in structural pathways that support functional circuits derived from medically healthy populations^30,59,60^. We have previously used white matter lesion network mapping to show that depression in MS is associated with lesions to the white matter structural backbone that supports a functional depression network derived from normative connectome data^30^. Here, we showed that lesions to the UF, the structural connection between key brain regions involved in stress circuitry, are associated with anxiety. Future studies that evaluate whether UF burden is associated with treatment outcomes are warranted.

### Generalizability and Scalability

To enhance the scalability of our study, we introduced an EMR phenotyping framework for assessing anxiety symptom severity that leverages known coding practices in working physicians and does not require prospective data collection. However, it is well-known that there are limitations to using the electronic medical record for psychiatric phenotyping^61^. First, anxiety is often underdiagnosed in medical populations^34,62^. As a result, an ICD-10 code for an anxiety disorder likely indicates the presence of anxiety symptoms, whereas the absence is not a confirmation that the patient is psychiatrically asymptomatic^33^. Additionally, many medications used for the treatment of anxiety are also approved for the treatment of depression, creating a further challenge in describing anxiety severity phenotypes without prospective assessments^38,57,63^.

We attempted to address these limitations by combining ICD-10 diagnoses and medication lists to stratify our anxiety groups, and included a mood screen to define our psychiatrically healthy comparators. However, this required making assumptions about how anxiety severity burden might manifest in provider diagnostic and prescribing behavior, which we cannot verify in our retrospective study. Though it is possible that the MS+severeA group could represent adequately treated patients with reduced anxiety symptom burden, our PROMIS scores confirmed that our phenotyping captured expected patterns described in prospectively acquired data samples, where worse anxiety was associated with worse functioning^64^. We also cannot exclude the possibility that comorbid symptoms such as pain or depression contributed to medication prescriptions in MS+mildA and MS+severeA, though the mean PHQ2 in all anxiety groups was less than 3, the threshold for a positive depression screen^65^. Psychiatric disease stratification using multi-level EMR-phenotyping with external validation may hold promise for EMR research across other psychiatric illnesses, medical populations, and health systems and more broadly.

### Limitations

There are several limitations to our study. First, our analysis is based on a lifetime history of anxiety rather than anxiety at the time of the scan. We attempted to mitigate this by validating our anxiety severity groups with PROMIS scores and time-linking them to their most proximal MRI, but they do not reflect psychiatric diagnoses, and the sample is small. Expanded Disability Status Scale (EDSS) scores, a measure of physical disability that is often collected in MS clinical trials but is not part of routine care^66,67^, were not available for our participants, though our PROMIS scores do quantify physical disability. Prospective studies that combine validated psychiatric symptom ratings with MS disability assessments and relate them to lesion burden would be informative.

We evaluated the UF in our anxiety analyses based on its anatomic connections with the OFC and amygdala, though UF variability has also been associated with language^68,69^ and memory^23^. While our sensitivity analyses suggest that the relationship between UF and anxiety may be specific, studies relating lesion burden to prospectively collected neurocognitive assessments are required^70^. Finally, our analyses focused on structural lesions without differentiating between lesion activity. Future studies that prospectively compare new lesions to anxiety measures are warranted.

## Conclusion

In this retrospective study, we explored the relationship between MS and anxiety. We developed a multi-step EMR phenotyping method to define anxiety groups and identified associations between anxiety diagnosis and severity to UF lesion burden. We showed that higher UF lesion burden was associated with an anxiety diagnosis, and worsening anxiety severity was associated with greater UF burden. This approach holds promise for understanding both anxiety in MS and the role of abnormalities in white matter as a mechanism for anxiety more broadly.

## Data Availability

Patient data are HIPAA-protected and cannot be shared publicly. Code and instructions for replicating all analyses can be found at: https://baller-lab.github.io/msanxiety/.

## Author Contributions

Drs. Baller, Satterthwaite, and Shinohara had full access to all the data in the study and take responsibility for the integrity of the data and the accuracy of the data analysis.

*Concept and design:* Baller, Schindler, Cieslak, Satterthwaite, Shinohara

*Acquisition, analysis, or interpretation of data:* Baller, Luo, Schindler, Cooper, Pecsok, Cieslak, Martin, Bar-Or, Perrone, Reid, Spangler, Satterthwaite, Shinohara.

*Drafting of the manuscript:* Baller, Satterthwaite, Shinohara.

*Critical revision of the manuscript for important intellectual content:* Baller, Luo, Cooper, Schindler, Cieslak, Satterthwaite, Shinohara.

*Statistical analysis:* Baller, Luo.

*Obtained funding:* Baller, Satterthwaite, Shinohara.

*Administrative, technical, or material support:* Cooper, Elahi, Reid

*Supervision:* Satterthwaite, Shinohara

This work was supported by grants from the National Institute of Mental Health (K23MH133118 to EBB; R01MH112847 to TDS and RTS; R01MH120482 and R01MH113550 to TDS; R01MH123550 to RTS), Brain and Behavior Research Foundation (NARSAD Young Investigator Award #31319 to EBB), the National Institute for Neurological Disorder and Stroke (R01NS085211 and R01NS112274 to RTS), and the National Multiple Sclerosis Society. Additional support was provided by the Penn-CHOP Lifespan Brain Institute.

