## Supplementary Tables for "Association of Anxiety with Uncinate Fasciculus Lesion Burden in Multiple Sclerosis"

Supplement

Table of Contents

| Supplemental Material |  | Page # |
| --- | --- | --- |
| 1. **Supplementary Table 1.** Medications used in the treatment of anxiety. |  | 2-3 |
| 2. **Supplementary Table 2.** Medications used in the treatment of multiple sclerosis. |  | 4-5 |
| 3. **Supplementary Table 3.** Demographics of sample with Patient-Reported Outcomes Measurement Information System (PROMIS) scores. |  | 6-7 |
| 4. **Supplementary Table 4.** Post hoc analyses of PROMIS summary measures. |  | 8 |

**Supplementary Table 1. Medications used in the treatment of anxiety.** Brand and generic medications that are FDA-approved for the treatment of anxiety, as well as medications commonly used off-label for anxiety (e.g. gabapentin, propranolol), are included^1^. Antipsychotic medications were excluded. NaSSA, noradrenergic and specific serotonergic antidepressant; SNRI, serotonin-norepinephrine reuptake inhibitor; SSRI, selective serotonin reuptake inhibitor, TCA, tricylic antidepressant.

|  | **Medication** | **Class** |
| --- | --- | --- |
| 1 | Gabapentin | Anticonvulsant |
| 2 | Lyrica | Anticonvulsant |
| 3 | Neurontin | Anticonvulsant |
| 4 | Pregabalin | Anticonvulsant |
| 5 | Atarax | Antihistamine |
| 6 | Hydroxyzine | Antihistamine |
| 7 | Vistaril | Antihistamine |
| 8 | Buspar | Anxiolytic |
| 9 | Buspirone | Anxiolytic |
| 10 | Alprazolam | Benzodiazepine |
| 11 | Ativan | Benzodiazepine |
| 12 | Chlordiazepoxide | Benzodiazepine |
| 13 | Clonazepam | Benzodiazepine |
| 14 | Diazepam | Benzodiazepine |
| 15 | Klonopin | Benzodiazepine |
| 16 | Librium | Benzodiazepine |
| 17 | Lorazepam | Benzodiazepine |
| 18 | Oxazepam | Benzodiazepine |
| 19 | Serax | Benzodiazepine |
| 20 | Valium | Benzodiazepine |
| 21 | Xanax | Benzodiazepine |
| 22 | Inderal | Beta-blocker |
| 23 | Propranolol | Beta-blocker |
| 24 | Mirtazapine | NaSSA |
| 25 | Remeron | NaSSA |
| 26 | Cymbalta | SNRI |
| 27 | Duloxetine | SNRI |
| 28 | Effexor | SNRI |
| 29 | Venlafaxine | SNRI |
| 30 | Celexa | SSRI |
| 31 | Citalopram | SSRI |
| 32 | Escitalopram | SSRI |
| 33 | Fluoxetine | SSRI |
| 34 | Lexapro | SSRI |
| 35 | Paroxetine | SSRI |
| 36 | Paxil | SSRI |
| 37 | Prozac | SSRI |
| 38 | Sertraline | SSRI |
| 39 | Zoloft | SSRI |
| 40 | Amitriptyline | TCA |
| 41 | Elavil | TCA |
| 42 | Nortriptyline | TCA |
| 43 | Pamelor | TCA |

**Supplementary Table 2. Medications used in the treatment of multiple sclerosis.** Brand and generic medications that are used for the treatment of multiple sclerosis, as well as their mechanism^2^. Anti-CD20, medications that deplete cells with CD20 receptor; DHODH-Inhibitor, inhibitor of dihydroorotate dehydrogenase; MBP, myelin basic protein; NRF-2, nuclear factor erythroid 2-related factor 2; S1P Agonist, sphingosine-1-phosphate. Medications listed are represented in our sample.

|  | **Medication** | **Mechanism** |
| --- | --- | --- |
| 1 | Natalizumab | A4 integrin-Inhibitor |
| 2 | Tysabri | A4 integrin-Inhibitor |
| 3 | Briumvi | Anti-CD20 |
| 4 | Kesimpta | Anti-CD20 |
| 5 | Ocrelizumab | Anti-CD20 |
| 6 | Ocrevus | Anti-CD20 |
| 7 | Ofatumumab | Anti-CD20 |
| 8 | Rituxan | Anti-CD20 |
| 9 | Rituximab | Anti-CD20 |
| 10 | Ublituximab | Anti-CD20 |
| 11 | Cladribine | Anti-Metabolite |
| 12 | Mavenclad | Anti-Metabolite |
| 13 | Aubagio | DHODH Inhibitor |
| 14 | Teriflunomide | DHODH Inhibitor |
| 15 | Avonex | Interferon |
| 16 | Avonex Pen | Interferon |
| 17 | Betaseron | Interferon |
| 18 | Extavia | Interferon |
| 19 | Interferons | Interferon |
| 20 | Plegridy | Interferon |
| 21 | Rebif | Interferon |
| 22 | Copaxone | MBP Analog |
| 23 | Glatiramer acetate | MBP Analog |
| 24 | Glatopa | MBP Analog |
| 25 | Fumaderm | Nrf-2 |
| 26 | Fumaric acid esters | Nrf-2 |
| 27 | Tecfidera | Nrf-2 |
| 28 | Gilenya | S1P Agonist |
| 29 | Fingolimod | S1P Agonist |
| 30 | Deltasone | Steroid |
| 31 | Medrol | Steroid |
| 32 | Methylprednisolone | Steroid |
| 33 | Prednisone | Steroid |
| 34 | Solu-Medrol | Steroid |
| 35 | Sterapred | Steroid |

**Supplementary Table 3. Demographics of sample with Patient-Reported Outcomes Measurement Information System (PROMIS) scores.** All demographic variables were extracted from the electronic medical record. Race is patient-reported. Lower scores represent more impairment. P-values reflect ANOVA tests in continuous data (age, Patient Health Questionairre-2, PROMIS scores) and ꭓ^2^ for categorical variables (sex, race, and MS medication). MS, multiple sclerosis; SD, standard deviation; MS+noA, MS without anxiety; MS+mildA, MS with mild anxiety (anxiety diagnosis *or* anxiolytic medication); MS+severeA, MS with severe anxiety (anxiety diagnosis *and* anxiolytic medication); anti-CD20, medications that deplete CD20 B-cells (**Supplementary Table 2)**.

|  | **MS+noA** | | | **MS+mildA** | **MS+severeA** | **P**  *Anxiety Diagnosis* | **P**  *Anxiety Severity* |
| --- | --- | --- | --- | --- | --- | --- | --- |
| n | | 12 | 45 | | 7 |  |  |
| Age (mean [SD]) | | 50.08 [10.30] | 49.76 [10.68] | | 46.29 [8.04] | N.S. | N.S. |
| Sex (% Male) | | 3 (25.0%) | 7 (15.6%) | | 0 (0%) | N.S. | N.S. |
| Race | |  |  | |  | N.S. | N.S. |
| Black or African American | | 2 (16.7%) | 11 (24.4%) | | 2 (28.6%) |  |  |
| Unknown | | 0 (0%) | 1 (2.2%) | | 0 (0%) |  |  |
| White | | 10 (83.3%) | 33 (73.3%) | | 5 (71.4%) |  |  |
| MS Medication | |  |  | |  |  |  |
| On Anti-CD20 | | 2 (16.7%) | 2 (4.4%) | | 4 (57.1%) | N.S. | <0.001 |
| On Interferon | | 0 (0%) | 0 (0%) | | 0 (0%) | N.S. | N.S. |
| On Steroids | | 0 (0%) | 0 (0%) | | 1 (14.3%) | N.S. | 0.016 |
| Patient Health Questionnaire-2 (mean [SD]) | | 0.0 [0.0] | 1.00 [0.85] | | 2.00 [2.08] | 0.003 | 0.002 |
| Quality of Life (mean [SD]) | | 4.25 [0.62] | 2.93 [1.01] | | 2.57 [0.98] | <0.001 | <0.001 |
| Physical Health (mean [SD]) | | 3.17 [0.58] | 2.47 [0.81] | | 2.29 [0.95] | 0.021 | 0.019 |
| Mental Health and Mood (mean [SD]) | | 3.92 [0.79] | 3.40 [0.89] | | 2.86 [1.21] | 0.034 | N.S. |
| Social Activities Satisfaction (mean [SD]) | | 4 [0.74] | 2.93 [1.27] | | 2.71 [1.50] | 0.022 | 0.022 |
| Carrying Out Social Activities (mean [SD]) | | 4 [0.85] | 3.11 [1.23] | | 2.29 [1.38] | 0.004 | 0.010 |
| Carrying Out Physical Activities (mean [SD]) | | 3.75 [0.87] | 3.16 [1.04] | | 3 [1.00] | N.S. | N.S. |
| Emotional Problems  (mean [SD]) | | 4.08 [0.51] | 3.09 [1.10] | | 2.71 [1.11] | 0.002 | 0.006 |
| Fatigue Average  (mean [SD]) | | 3.75 [0.75] | 3.16 [1.19] | | 3.14 [1.35] | N.S. | N.S. |
| Post-Op PROMIS Physical Score (mean [SD]) | | 14.42 [2.15] | 11.67 [2.80] | | 11.57 [3.36] | 0.037 | 0.011 |
| Post-Op PROMIS Mental Score (mean [SD]) | | 16.25 [2.01] | 12.36 [3.16] | | 10.86 [3.58] | 0.001 | <0.001 |

**Supplementary Table 4.** **Post hoc analyses of PROMIS summary measures.** Pairwise contrasts are reported as P-values with corresponding Cohen’s d. Multiple comparisons were accounted for by controlling the False Discovery Rate (Q < 0.05). MS+noA, MS without anxiety; MS+mildA, MS with mild anxiety (anxiety diagnosis *or* anxiolytic medication); MS+severeA, MS with severe anxiety (anxiety diagnosis *and* anxiolytic medication).

|  | **T** | **P*_fdr_*** | **Cohen’s *d*** |
| --- | --- | --- | --- |
| Physical vs Emotional Functioning |  |  |  |
| MS+noA | 0.68 | >0.05 (N.S) | - |
| MS+mildA | 2.97 | 0.015 | -0.38 |
| MS+severeA | 2.88 | 0.047 | -0.69 |
| Emotional Functioning MS+severeA vs MS+noA | 3.68 | 0.015 | -1.87 |
| Physical Functioning MS+severeA vs MS+noA | 2.00 | >0.05 (N.S) | - |
